# Early chains of transmission of COVID-19 in France

**DOI:** 10.1101/2020.11.17.20232264

**Authors:** Juliette Paireau, Alexandra Mailles, Catherine Eisenauher, Franck de Laval, François Delon, Paolo Bosetti, Henrik Salje, Valérie Pontiès, Simon Cauchemez

## Abstract

**Introduction:** SARS-CoV-2, which causes COVID-19, has spread rapidly across the world. A dedicated surveillance system was implemented in France in January 2020 to improve early detection of cases and their contacts and limit secondary transmission. Our objective was to use contact-tracing data collected during this initial phase of the epidemic to better characterize SARS-CoV-2 transmission.

**Methods:** We analysed data collected during contact tracing and retrospective epidemiological investigations in France from 24 January to 30 March 2020. We assessed the secondary clinical attack rate and characterized the risk of a contact becoming a case. We described chains of transmission and estimated key parameters of spread.

**Results:** Over the study period, 6,082 contacts of 735 confirmed cases were traced. The overall secondary clinical attack rate was 4.1% (95%CI 3.6-4.6) and increased with age of the index case and of the contact. Family contacts were at higher risk of becoming cases (adjusted odds ratio 2.1 (95%CI 1.4-3.0)) while nosocomial contacts were at lower risk (adjusted odds ratio 0.3 (95%CI 0.1-0.7)), compared to coworkers/friends. We identified 328 infector/infectee pairs, 49% of which were family members. The distribution of secondary cases was highly over-dispersed with 80% of secondary cases being caused by 10% of cases. The mean serial interval was 5.1 days (interquartile range 2-8 days) in contact-tracing pairs where late transmission events may be censored, and 6.8 (3-8) days in pairs investigated retrospectively.

**Conclusion:** This study contributes to improving our knowledge of SARS-CoV-2 transmission, such as the importance of superspreading events. Contact-tracing data are challenging to collect but are key to better understand emerging pathogens.

**Funding statement:** This work was supported by the LabEx “Integrative Biology of Emerging Infectious Diseases (IBEID)” (Grant Number ANR-10-LABX-62-IBEID), Santé Publique France, the INCEPTION project (PIA/ANR-16-CONV-0005), and the European Union’s Horizon 2020 research and innovation program under grants 101003589 (RECOVER) and 874735 (VEO).

## Introduction

Coronavirus disease 2019 (COVID-19) emerged in Wuhan, China, in December 2019 and has since spread globally. The disease is caused by severe acute respiratory syndrome coronavirus 2 (SARS-CoV-2) and is transmitted from person to person mainly via small droplets produced by coughing, sneezing or talking. The rapid spread of the virus across the world has led to the implementation of unprecedented containment measures, with more than two billion individuals being confined at home. Following a fast rise in Intensive Care Unit (ICU) admissions, France went into lockdown on 17 March 2020 for seven weeks. As of 20 October 2020, 930,745 cases have been confirmed, including 142,654hospitalizations and 33,885 deaths (1).

In the early phase of the epidemic, France attempted to contain importations into its territory. On 10 January 2020, a dedicated surveillance system was implemented to allow early detection of cases and their contacts, limit secondary transmission and slow the spread of the virus. Upon detection of a COVID-19 case, contact tracing was initiated and a follow-up procedure was implemented. The first three COVID-19 cases were detected on 24 January 2020 in travellers returning from Wuhan (2).

Contact tracing is an essential tool to control epidemics and has proven efficient in the past (3,4). It can also contribute to improving our knowledge on transmission dynamic and natural history of emerging pathogens such as SARS-CoV-2. Contact-tracing studies can provide estimates of secondary attack rates, explore risk factors of infection among contacts, and assess the relative contributions of different types of contact to transmission, which is key to defining efficient control strategies (5–11).

Here, we analysed the detailed data collected during contact tracing and retrospective epidemiological investigations, in the early phase of the epidemic in France, from 24 January 2020 until 30 March 2020, two weeks after lockdown. The objectives of our study were: (1) to assess the secondary clinical attack rate and identify the factors associated with the risk of a contact becoming a case and (2) to describe chains of transmission and estimate key parameters of spread.

## Methods

### Surveillance system

The individual-based surveillance system aiming at detecting cases was implemented on 10 January 2020 (see (2) for a detailed description and Text S1 for case definitions).

Possible cases were isolated in one of the COVID-19 reference hospitals whenever possible, or at home, and interviewed using a standardised questionnaire that collected information on sociodemographics, clinical characteristics, and history of exposure (including potential infectors). Data were entered into a secure web-based application (Go.Data©, WHO). Respiratory samples were taken from all possible cases and tested for SARS-CoV-2 using RT-PCR. A confirmed case was defined as a possible case with a positive RT-PCR.

From 14 March 2020, the system was replaced by a population-level approach, which was more adapted to the growing size of the epidemic. However, individual-based surveillance continued for a few more days in less affected regions.

### Contact tracing

Confirmed cases were kept in isolation and interviewed about any contacts that occurred during the time they were symptomatic and in the day prior to symptom onset. Identified contacts were classified into 3 levels of exposure risk: negligible, low, or moderate/high risk, depending on their type of exposure (Table S3). Only contacts who developed symptoms compatible with COVID-19 were tested for SARS-CoV-2 by RT-PCR. If the test was positive, they were considered secondary confirmed cases and their contacts were then traced in the same fashion as a primary case. Following the procedures, only contacts with low or moderate/high risk were followed up (Table S3). In practice, due to a quick overload of investigation teams in the exponential phase of the epidemic, most contacts (97%) who were identified and entered into the database were moderate/high-risk contacts. In regions heavily affected by the epidemic, some cases had their contacts traced but not entered into the web-based application. Therefore, the database is not exhaustive and represents a sample of all the contact tracing efforts. In some of these regions, contact tracing became too difficult to conduct and was stopped before 14 March.

### Retrospective epidemiological investigations

In addition to the chains of transmission established through prospective contact tracing, some infector/infectee pairs were reconstituted retrospectively during epidemiological investigations. This was especially the case in the Oise department in northern France, where a large cluster of COVID-19 was detected in February (12). A thorough investigation, aiming at reconstructing the chains of transmission underlying the cluster and understanding the rapid spread of the pathogen, was conducted in this department (Text S2).

### Ethics statement

The investigations were carried out in accordance with the General Data Protection Regulation (Regulation (EU) 2016/679 and Directive 95/46/EC) and French data protection law (Law 78–17 on 06/01/1978 and Décret 2019–536 on 29/05/2019).

### Statistical analysis

We first analysed contact-tracing data for confirmed cases (simply called “cases” thereafter) and their contacts entered into the database between 24 January, 2020 and 30 March, 2020. We define as an “index case” a case whose detection initiated an investigation of its contacts through contact tracing.

We described contact patterns between index cases and their traced contacts by constructing the corresponding contact matrix. We compared it to the age-specific contact matrix for the French population obtained from the COMES-F study performed in 2012 (13). We analysed the differences in the age-specific mixing patterns with a linear regression.

We estimated the secondary clinical attack rate, defined as the proportion of symptomatic cases among the contacts of an index case (14). We investigated the factors associated with the risk of a contact becoming a case (i.e. developing symptoms and testing positive) using multivariable logistic regression (Text S3).

Finally, we analysed all infector/infectee pairs, using pairs identified through prospective contact tracing (pairs between an index case and a contact who became a case) and pairs identified through retrospective epidemiological investigations in Oise. We estimated the number of secondary cases generated by each case and the serial interval (Text S4).

Distributions were compared using Wilcoxon-Mann-Whitney test and proportions were compared using chi-square test. Results with p-value <0.05 were considered significant. All analyses were performed in R software (15).

## Results

### Description of contact-tracing data

Between 24 January and 30 March 2020, 6,082 contacts (corresponding to 6,028 unique individuals) of 735 confirmed cases were traced and entered into the database. The median age of the index cases was 50 years (interquartile range (IQR) 36-65) and 52% (384/734) were female. The median age of the contacts was 38 years (IQR 21-55) and 56% (3123/5593) were female.

Cases had 8.3 contacts traced on average (median 4, maximum 146). We observed non-random contact patterns by age, with index cases tending to have a higher number of traced contacts of similar age (Pearson correlation coefficient r = 0.32, p<0.01) (Figure 1A). These patterns were consistent with age-specific contacts measured prior to the epidemic in the French general population (Pearson r = 0.80, p<0.01) (Figure 1B and Figure S1). The transmission pairs followed a similar assortative pattern by age (Pearson r = 0.29, p<0.01) (Figure 1C).

**Figure 1:**
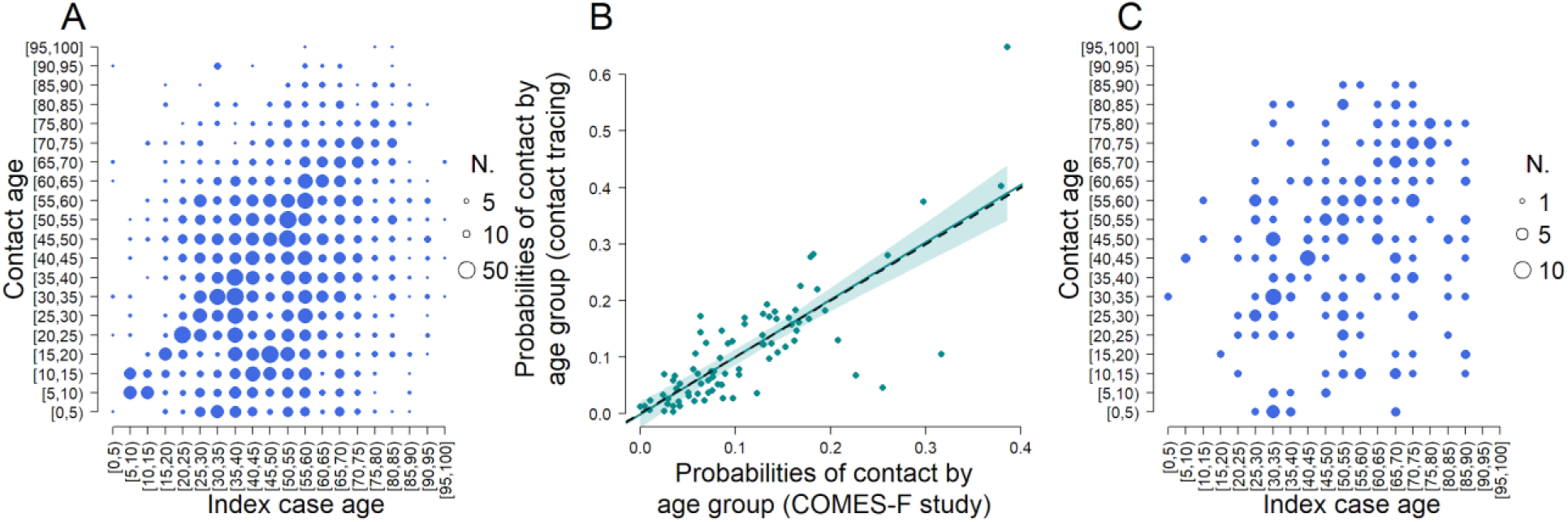
Distribution of contacts and cases by age. (A) Number of contacts in each index case/contact combination. (B) Correlation between contacts patterns in the contact tracing and contacts patterns in the general population (see also Figure S1). (C) Number of secondary cases in each index case/contact combination.

The 735 confirmed cases that had their contacts traced and entered into the database represented 5% (735/14,400) of the total number of confirmed cases over the study period. This proportion decreased over time, from 31% at the beginning of the epidemic to 2% in weeks 12-13 (week 11 was the last week of the individual based-surveillance at the national level) (Figure 2A). The proportion of confirmed cases that had their contacts traced and entered into the database also varied by region (Figure 2B). The most heavily affected regions (Ile-de-France (IDF) and Grand Est (GES)), representing 54% of confirmed cases, had the lowest proportion of traced cases (1%).

**Figure 2:**
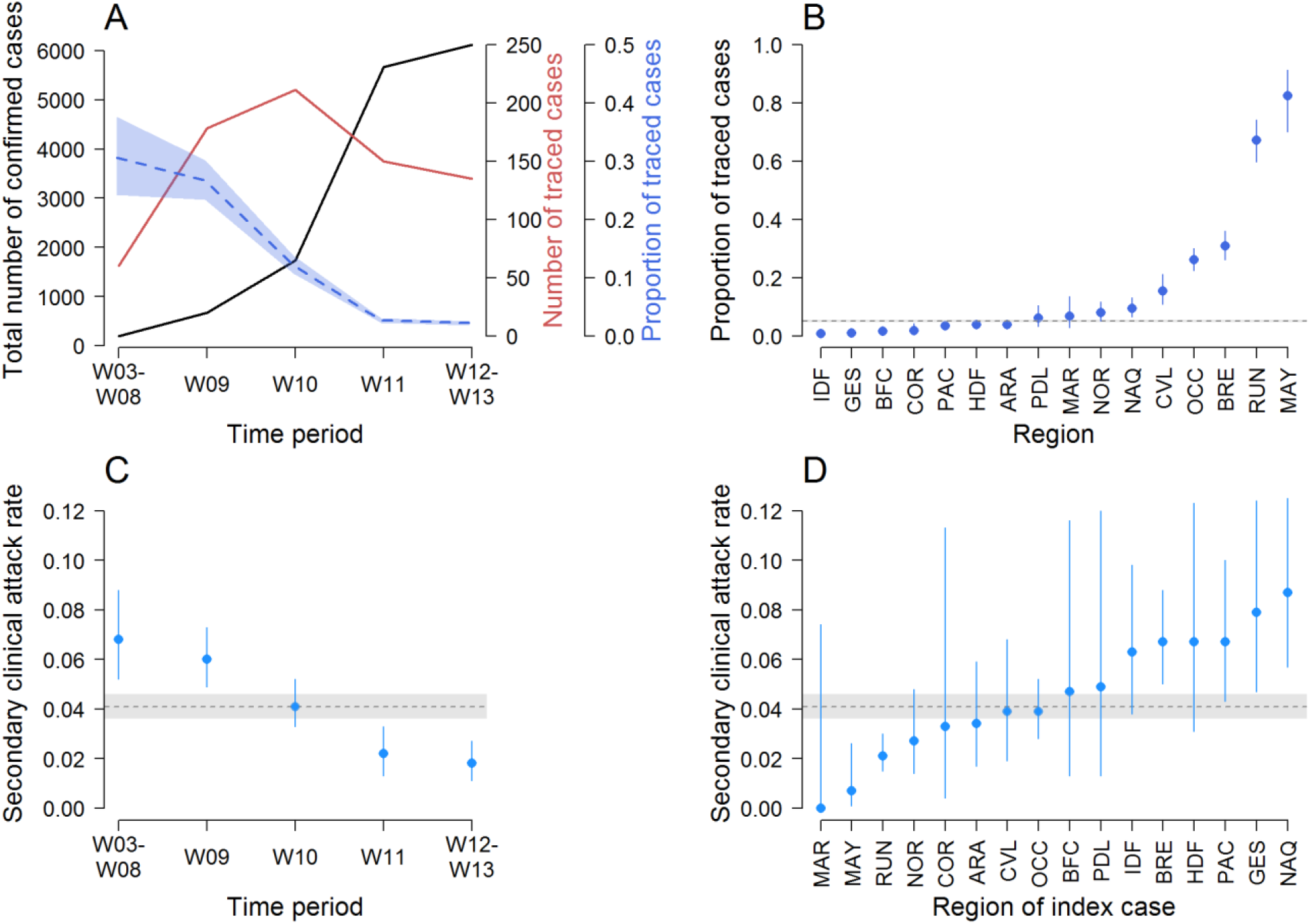
Description of contact-tracing data over time and regions. (A) Number of confirmed and traced cases, and proportion of confirmed cases that had their contacts traced and entered into the database, by time period (onset weeks). Week 11 was the last week of the individual-based surveillance at the national level. (B) Proportion of confirmed cases that had their contacts traced and entered into the database by region. (C) Secondary clinical attack rate over time. (D) Secondary clinical attack rate by region. The horizontal dashed lines indicate the national values over the whole time period. Shaded areas and vertical bars represent 95% confidence intervals.

### Secondary clinical attack rate

Among the 6,028 contacts traced and entered in the database, 248 became secondary cases, representing an overall secondary clinical attack rate of 4.1% (95%CI 3.6-4.6). The secondary clinical attack rate among contacts of secondary cases was 2.3% (95%CI 1.4-3.6).

The secondary clinical attack rate decreased over time, from 6.8% at the beginning of the epidemic to 1.8% in weeks 12-13 (Figure 2C). It also varied by region, from 0% to 8.7% (Figure 2D). The secondary clinical attack rate increased with the age of the contact, ranging from 4.7% (95%CI 3.2-6.7) for contacts aged 0-14 years to 12.2% (95%CI 8.0-17.7) for contacts who were 75 or older, and was not different by gender of the contact (Figure 3A). The secondary clinical attack rate also increased with the age of the index case, from 2.0% (95%CI 0.7-4.7) for 0-14 years old to 6.2% (95%CI 4.3-8.7) for 75+, although we should note the very small number of young index cases (N=15) (Figure 3B). It did not vary with the gender of the index case, except in individuals 75 or older for which it was higher for male (9.8% (95%CI 6.7-13.7)) than female index cases (0.5% (95%CI 0.0-2.9)). The secondary clinical attack rate was the highest among family members (7.9% (95%CI 6.6-9.3)), followed by coworkers/friends (3.4% (95%CI 2.5-4.4)), those travelling with a case (3.4% (95%CI 1.9-5.4)), and nosocomial contacts (1.1% (95%CI 0.4-2.3)) (Figure 3C).

**Figure 3:**
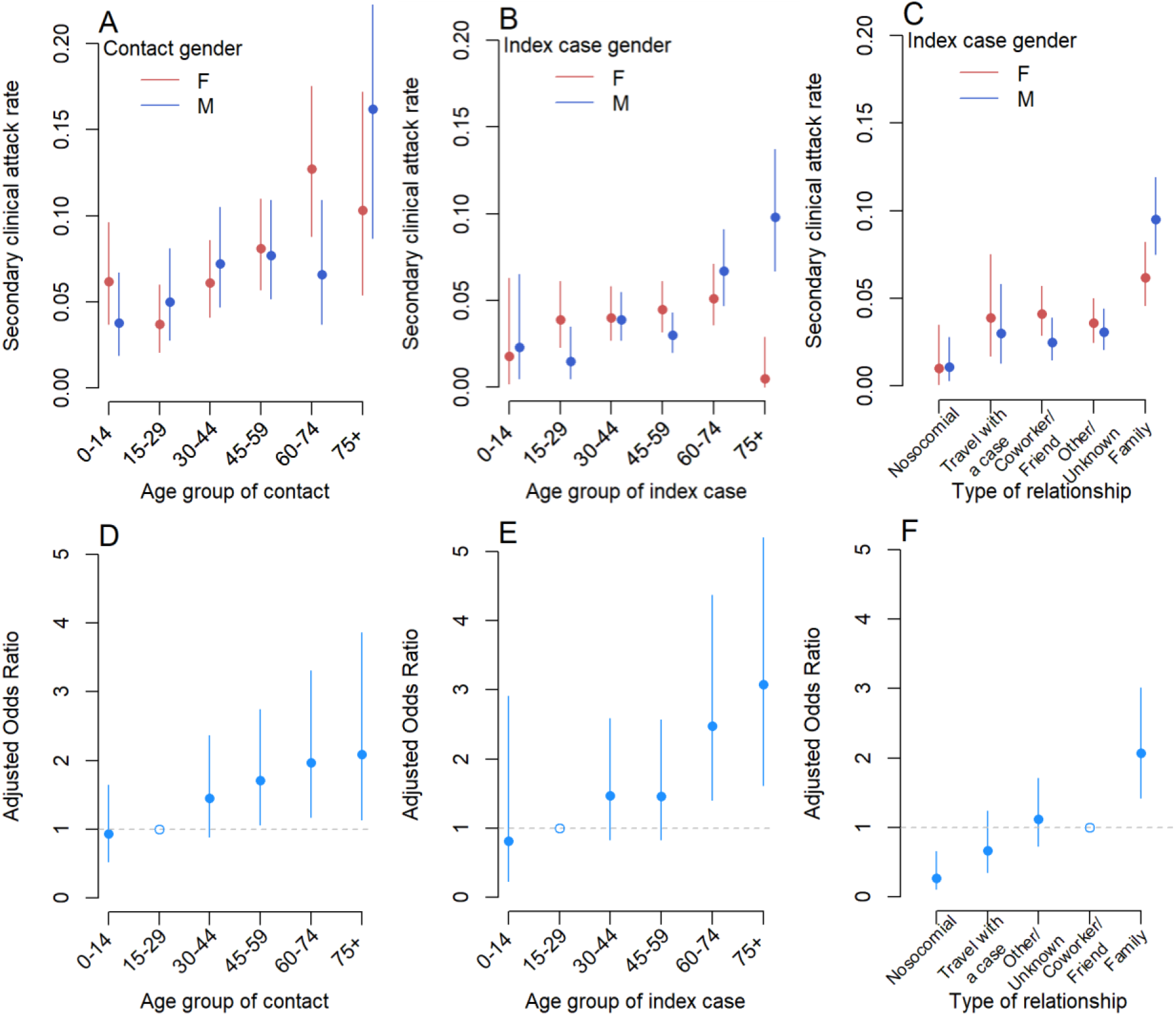
Secondary clinical attack rate and factors associated with the risk of a contact becoming a case. Secondary clinical attack rate according to the age and gender of the contact (A), the age and gender of the index case (B) and the type of relationship between the contact and the index case and gender of the index case (C). Adjusted odds ratios of the association between contact becoming a case and contact age (reference: 15-29 years old) (D), index case age (reference: 15-29 years old) (E) and type of relationship (reference coworker/friend) (F).

### Factors associated with the risk of a contact becoming a case

In univariable analysis, the gender of the index case and of the contact were not associated with the risk of a contact becoming a case. Therefore, only the type of relationship, the age of the contact and of the index case were considered for the multivariable model. All three of these risk factors remained significantly associated with the risk of a contact becoming a case in the multivariable analysis. The odds of becoming a case were highest for contacts aged 45-59 years (adjusted odds ratio 1.7 (95%CI 1.212.7), 60-74 years (adjusted odds ratio 2.0 (95%CI 1.2-3.3)), and older than 75 years old (adjusted odds ratio 2.1 (95%CI 1.1-3.9)), compared to the reference group 15-29 years old (Figure 3D). The odds were higher for contacts whose index case was 60-74 (adjusted odds ratio 2.5 (95%CI 1.4-4.4)) or older than 75 years old (adjusted odds ratio 3.1 (95%CI 1.6-5.8)), compared to the reference group 15-29 years old (Figure 3E). Contacts of index cases aged less than 15 years had a similar risk than contacts of index cases aged 15-29 years (adjusted odds ratio 0.8 (95%CI 0.2-2.9)). We found no significant interaction between the age of the index case and that of the contact. Family contacts were at higher risk of becoming cases (adjusted odds ratio 2.1 (95%CI 1.4-3.0)) and nosocomial contacts were at lower risk (adjusted odds ratio 0.3 (95%CI 0.1-0.7)), compared to coworkers/friends (Figure 3F). In the sensitivity analyses accounting for regional and temporal differences in data collection, and restricting the data to moderate/high-risk contacts, the estimates were not substantially modified compared to the baseline model (Figures S2 and S3). The sensitivity analysis with inclusion of contacts with multiple index cases by random assignment of a single index case also showed results consistent with the baseline analysis (Figure S4).

### Chains of transmission

Overall, 328 connections between cases were identified, representing plausible transmission events between an infector and an infectee (Figure 4). Among those, 259 infector/infectee pairs were identified through contact tracing and 69 reconstituted retrospectively through epidemiological investigations in Oise. These pairs involved 418 individuals, including 154 infectors. In total, 109 unique transmission chains with at least 2 cases were identified with a median size of 2 individuals and a mean size of 3.8 individuals (Figure 5A). The largest chain included 39 cases and spanned 5 generations (Figure 5B).

**Figure 4:**
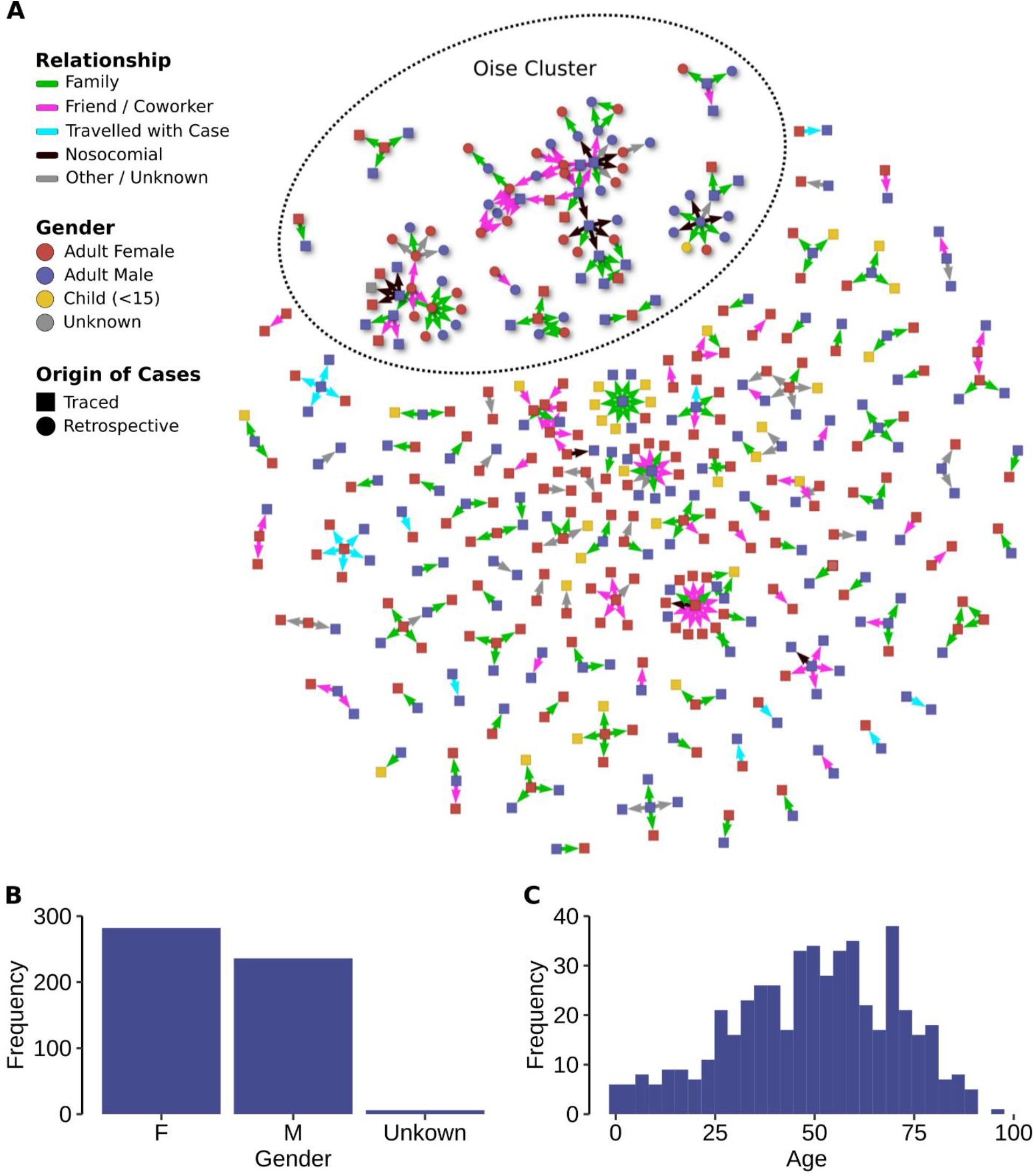
Chains of transmission. (A) Observed transmission chains. Only confirmed cases/contacts involved in a chain with at least one confirmed case are represented. (B) Distribution of sex. (C) Distribution of age.

**Figure 5:**
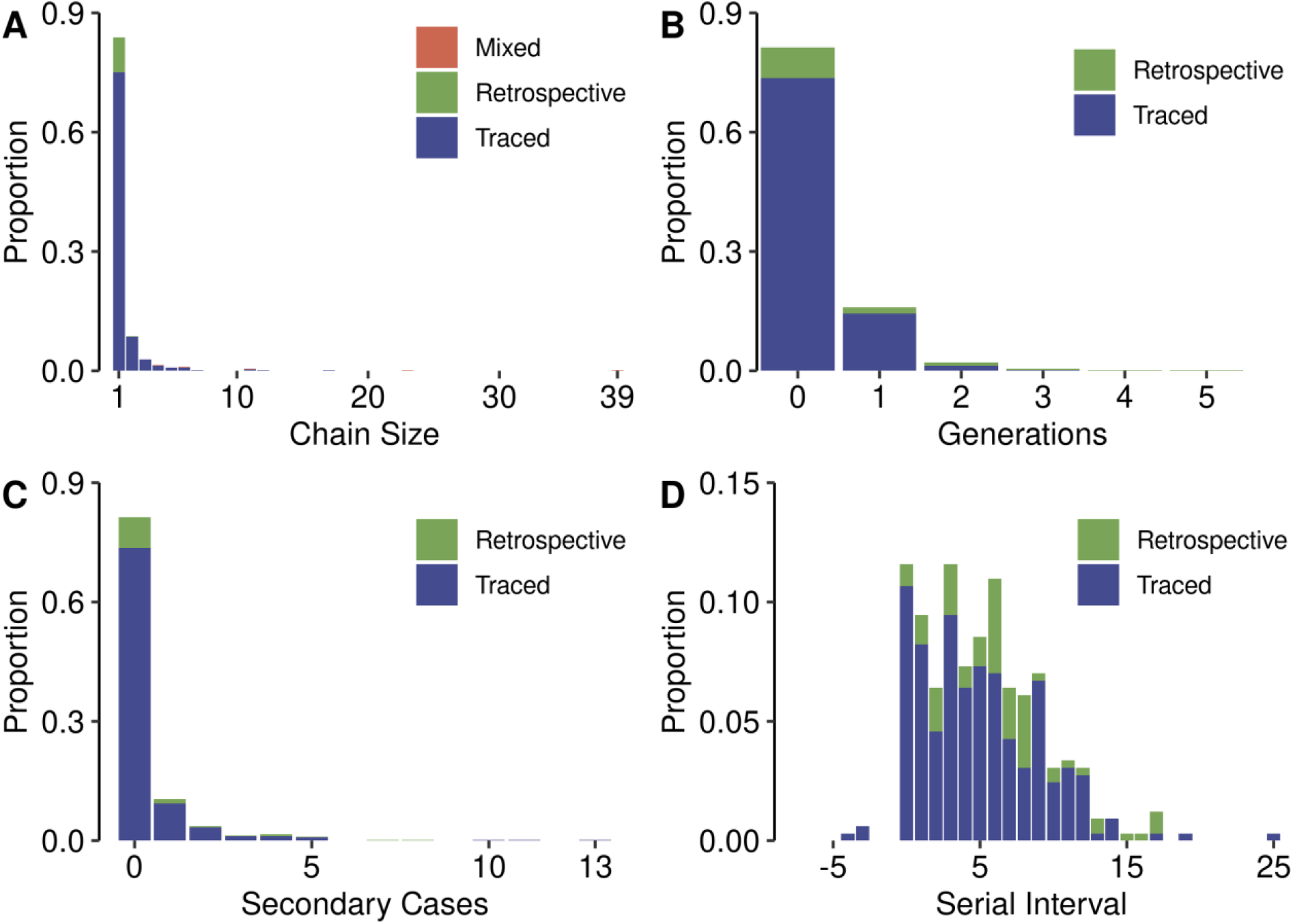
Summary Statistics. (A) Distribution of chain size. (B) Distribution of number of onward generations of cases resulting from a case (cases with no infected contacts have zero onward generations). (C) Distribution of number of secondary cases arising from a case. Note that for contact tracing, this distribution represents all index cases, including those who had no identified transmission chain and are therefore not in the transmission network panel (Figure 4A). For retrospective investigations, this distribution represents all infectors and infectees, including terminal infectees (cases with no identified onward transmission). (D) Distribution of serial interval in days. In each panel, we indicate the origin of the chains/cases, either from contact-tracing data (“traced”), from retrospective epidemiological investigations (“retrospective”), or both (“mixed”).

In Oise, the first two cases were identified on 25 February 2020, with onset dates on 10 February. Epidemiological investigations showed that these two cases were part of a larger cluster, with several large chains of transmission occurring in a military support facility, a secondary school and a high school. Several transmission events were also associated with nosocomial settings such as two hospitals, a clinic, and a general practice. In total, nine transmission chains were reconstructed, including the two largest chains of 39 and 23 cases, spanning 5 and 4 generations respectively. Some cases belonging to these chains also gave rise to secondary cases in other departments. A few other transmission events (smaller chains in Figure 4A) were identified in this department during the investigation period but could not be formally linked to the large chains. In total, 97 infector/infectee pairs could be established in this cluster, including 69 through retrospective investigations and 28 through prospective contact tracing.

Among pairs identified through contact tracing, the median age of the infectors was 51 years (interquartile range (IQR) 37-67), 3% (4/127) were children (under 15 years old) and 51% (67/131) were female. The median age of the infectees was 48 years (IQR 30-62), 12% (28/236) were children and 55% (137/247) were female. These characteristics were similar in the retrospective pairs, except that infectees were older than in contact tracing (median age 54 vs 48 years, p=0.01) (Table S4). In contact-tracing pairs, infectors and infectees had a family link in 52% of the pairs, 26% were coworkers or friends, 7% of the infectees had travelled with their infector, 3% of the pairs were associated to nosocomial transmission, and 12% of the relationships were labelled as “other/unknown”. In the retrospective pairs, the proportion of nosocomial transmission was higher than in contact tracing (14% vs 3%, p<0.001) (Table S4).

In contact tracing data, index cases had 0.3 (95%CI 0.3-0.4) detected secondary cases on average (Figure 5C). The distribution of secondary cases was highly over-dispersed, with 80% of detected secondary cases being caused by 10% of cases (negative binomial dispersion parameter 0.17; 95%CI 0.12-0.22). In retrospective investigations, the mean number of secondary cases was 0.9 (95%CI 0.5-1.5) and 80% of detected secondary cases were caused by 16% of cases (dispersion parameter 0.28; 95%CI 0.09-0.47). Six super-spreading events were observed with 3 infectors generating 7-8 secondary cases (identified through retrospective investigations) and 3 infectors generating 10-13 secondary cases (identified through contact tracing). These transmission events occurred in the workplace (10 cases), during a dinner between neighbours (6 cases), a family/religious gathering (10 cases), or included mixed types of relationships (family/nosocomial/coworker, 31 cases). Among the 328 reported serial intervals (Figure 5D), 3 (0.9%) were negative and 38 (12%) were null. The serial interval had an average of 5.1 days (median 5, IQR 2-8 days) when calculated on pairs from contact tracing, and an average of 6.8 days (median 6, IQR 3-8) when calculated on pairs from retrospective investigations.

## Discussion

Using data from contact-tracing and epidemiological investigations conducted during the initial phase of the COVID-19 epidemic in France, we were able to characterize the secondary clinical attack rate and the factors associated with the risk of a contact becoming a case among 6082 contacts of 735 index cases. We also analysed chains of transmission and estimated key parameters of spread among 328 infector/infectee pairs.

Overall, 4.1% of contacts identified and entered into the database became secondary confirmed cases. Since only symptomatic contacts were tested and a proportion of persons infected by SARS-CoV-2 do not develop symptoms (16,17), it is most likely that some asymptomatic infections were missed among contacts. The definition of a contact might vary between studies and impact estimates of secondary attack rate. Cheng *et al* found a lower secondary clinical attack rate of 0.7% (9), but the number of contacts identified per index case was much higher (27 on average, compared to 8 in our study) and may have included more low-risk contacts than in our study, in which most contacts identified and recorded in the database were moderate/high-risk contacts. Conversely, in studies where 5-10 contacts were identified per index case, the secondary attack rate (including asymptomatic patients) was 3.7-10.7% (5,6,8,11). We found the highest secondary clinical attack rate among family contacts (7.9%, including household and non-household members, which we could not distinguish), highlighting the substantial risks associated with SARS-CoV-2 transmission between close family members. Family contacts might also be easier to identify as they constitute a close and well defined population contrary to other types of contacts such as coworkers or friends who may be more difficult to define. In contact-tracing studies where asymptomatic patients were tested, the household secondary attack rate was found to be around 9-17% (5,7,8,10,11), but a recent study found attack rates as high as 53% (18). The secondary clinical attack rate was lower among nosocomial contacts, in agreement with other studies (6,8,9,11). We also found a lower secondary clinical attack rate among contacts of secondary cases compared to all cases, suggesting the positive impact of isolation measures.

In our study population, we found that the risk of becoming a case was more than twice higher for contacts older than 45 years compared to 15-29 years old. This might be because older individuals develop more severe symptoms and are thus more likely to be detected (19,20). Since contact tracing is triggered by the detection of a symptomatic case and most infected children appear to be asymptomatic or mildly symptomatic (21), only 15 index cases out of 735 were children in our study. Given this important selection bias and the small number of children, our data do not make it possible to robustly compare the infectiousness of children relatively to adults.

Interestingly, the age-specific contact patterns observed in our study before cases are isolated were very consistent with those obtained in a large-scale population survey conducted in France in 2012 (13). This suggests that the contact-tracing data are representative of contact patterns in the general population, and that the Béraud’s contact matrix is appropriate to model the early dynamic of the epidemic, before lock-down, as was for example done in Salje *et al* (19).

We found an average serial interval of 5.1 days in contact-tracing pairs, consistent with published estimates of 4 to 6 days obtained in similar contexts of case isolation (5,22–26). This must be considered a lower bound of what would happen in a situation without control measures as the isolation of cases has a truncating effect. Such truncation has been demonstrated by Bi *et al* in their observation that the serial interval increased with delays in isolating cases, from 3.6 days if the infected individual was isolated less than 3 days after infection, to 8 days if the infected individual was isolated on the third day after symptom onset or later (5). This is consistent with our estimate of 6.8 days for the serial interval in pairs of infectors/infectees that were identified retrospectively and for which case isolation was likely more limited/delayed than in contact traced pairs.

The mean number of secondary cases identified per index case was 0.3-0.9. These values are lower than the estimates for the reproduction number of SARS-CoV-2 in the absence of interventions which are around 2.5-3 (19). This can be explained by a combination of several factors: (1) the number of secondary cases could be reduced due to contact tracing and isolation measures, (2) asymptomatic infections are not observed in the study settings, (3) some symptomatic secondary cases might have been missed despite contact tracing, or not recorded in the database. Other studies conducted in a similar context obtained similar results, with 0.4-0.7 secondary cases identified per index case (5,7,27). We estimated the dispersion parameter of the secondary cases distribution to be around 0.15-0.30, indicating a high degree of superspreading. This result adds to the growing evidence that transmission of SARS-CoV-2 is highly over-dispersed (27–30). It has important implications for control efforts: interventions targeting settings where such superspreading events occur could substantially reduce overall transmission.

Our study has several limitations. Data collected during outbreaks are often noisy and incomplete due to the difficult conditions in which they are collected. Case definitions and protocols evolved during the study period to adapt to the changing epidemic situation and new knowledge about the virus and its transmission. More importantly, there are major practical challenges associated with contact tracing, in particular the difficulty of both identifying all potential contacts of an individual and then closely monitoring those contacts during the recommended follow-up period of 14 days, with limited human resources. We showed that the proportion of traced cases and the secondary clinical attack rate declined over time, and varied across regions. This likely reflects variations in data quality and completeness, rather than the true evolution of the epidemiological situation due to control measures. Indeed, contact tracing could not be scaled up to meet the exponentially growing burden during the early phase of the epidemic, and investigation teams were quickly overwhelmed with the influx of new cases. Moreover, the work overload was heterogeneous between regions, depending on the local epidemiological situation, and data may therefore vary in quality and consistency. The northeast quarter of France was the most severely affected (especially Ile-de-France and Grand Est regions), while the rest of the country was less overwhelmed (19). However, in a sensitivity analysis, our risk-factor estimates were robust when accounting for regional and temporal differences in data collection. Another difficulty was to collect data on contacts and cases in healthcare settings. Given that the priority for hospital staff was patients’ care, it is perfectly understandable that, with the influx of new patients, only a short amount of time was left for epidemiological investigations. As a consequence, a substantial proportion of contacts and secondary cases in hospital settings may have been missed, potentially leading to an under-estimation of the role of the hospital in SARS-CoV-2 transmission. It is interesting to observe that in contact-tracing data, nosocomial transmission was documented in 3% of the infector/infectee pairs, while it rose to 14% in retrospective pairs in Oise, where thorough investigations probably helped to better document nosocomial transmission. The contribution of health care settings to overall transmission seems highly variable across studies (6,8,9). Finally, the infector/infectee pairs were established by the investigators based on the known relationships between the index and secondary cases and the circumstances of their contact. However, these putative transmission events are not biologically proven and we cannot exclude that some cases have been exposed to other infected persons. In addition, since not all cases initiated an investigation of their contacts, the transmission chains that we observe are incomplete and their size underestimated.

In conclusion, this study contributes to improving our knowledge and understanding of COVID-19. Despite the immense efforts necessary to perform contact tracing during outbreaks, collecting and analysing this type of data is important to better understand disease transmission, and can help to define efficient control strategies.

## Data Availability

Access to the data must be granted by Sante Publique France to protect patient privacy.

## Acknowledgements

We are indebted to the work of public health staff in Santé publique France regional offices and headquarters, and Agences Régionales de Santé, who are involved in COVID-19 surveillance and greatly contributed to contact tracing, retrospective investigations, data collection and data management. We thank Sylvie Haeghebaert, Caroline Vanbockstael and the Service de Santé des Armées for contributing to retrospective investigations in Oise, and Daniel Lévy-Bruhl and Sibylle Bernard-Stoecklin for useful discussions. We thank all patients and contacts involved in the study, as well as the medical staff.

## Authors’ contributions

Juliette Paireau, Alexandra Mailles, Henrik Salje, Valérie Pontiès and Simon Cauchemez conceived and designed the analysis.

Alexandra Mailles, Franck de Laval, François Delon and Valérie Pontiès contributed to retrospective investigations and contact tracing, and collected the data.

Juliette Paireau, Catherine Eisenhauer and Paolo Bosetti performed the statistical analysis.

Juliette Paireau and Simon Cauchemez wrote the first draft of the manuscript. All authors contributed to the final manuscript.

