## Supplementary material for "Early chains of transmission of COVID-19 in France"

**Text S1: Case definition**

From 17 to 29 January 2020, a possible case was defined either as a patient with a severe acute lower respiratory infection requiring admission to hospital and with a history of travel to or residence in Wuhan, China in the 14 days before symptom onset, or a patient with an acute respiratory illness whatever the severity and with a history of at-risk exposure, mainly to a confirmed case (1). This definition slightly evolved during the study period, to adapt to the epidemiological situation. The detailed case definition used at the start of the epidemic, as well as the case definition in effect at the end of the study period, are available in Tables S1 and S2. Possible cases who tested negative were classified as excluded cases.

**Text S2: Retrospective investigations**

Cases were interviewed about the contacts they had during the 14 days prior to symptom onset: contacts with sick individuals, contacts with individuals returning from a trip in a risk zone, or participation to any kind of gathering during a prolonged time period. The contact individuals identified through these interviews were then reached to confirm their link with the case, and included as cases in the study if they had symptoms. The pairs were considered as infector/infectee pairs if their characteristics (time between exposure and symptom onset, incubation period, duration and nature of the contact…) were compatible with SARS-CoV-2 transmission based on current knowledge. Data collected through these investigations were entered in the same database as cases and contact-tracing data.

**Text S3: Multivariable logistic regression model**

We investigated the factors associated with the risk of a contact becoming a case (i.e. developing symptoms and testing positive) using multivariable logistic regression. The model considered associations with age (categorized in 15-year age groups with 15-29 years old as reference), gender and type of relationships. We defined five categories for the type of relationships: (1) family, (2) coworkers (including teachers/students), friends or acquaintance (including neighbours and people regularly sharing leisure, community or religious activities), (3) travel with a case (i.e. the index case and the contact travelled together or shared the same transportation), (4) nosocomial contact (contact in a hospital, a general practice or other healthcare facility), (5) other/unknown. We classified as nosocomial the relationships that were both in family and nosocomial categories, and as family the relationships that were both in family and travel categories, and as coworkers/friends the relationships that were both in coworkers/friends and travel categories. Variables with a p-value less than 0.20 in univariable analyses were included in a multivariable model and iteratively removed using backward selection until all variables in the model had p-values less than 0.05. We performed three sensitivity analyses to assess the robustness of our model. First, we included the region of the index case and the time period as a random effect, in order to assess how our estimates were modified when accounting for regional and temporal differences in contact tracing. Second, we restricted the analysis to moderate/high-risk contacts only. Third, fifty-three contacts had multiple index cases (52 had two index cases and one contact had three index cases). These contacts were removed from the baseline analysis. We evaluated the sensitivity of risk-factor estimates to inclusion of these contacts, with random assignment of a single index case.

**Text S4: Analysis of infector/infectee pairs**

We analysed all infector/infectee pairs, using pairs identified through prospective contact tracing (pairs between an index case and a contact who became a case) and pairs identified through retrospective epidemiological investigations in Oise. We described the transmission network in terms of chain size (number of connected nodes) and number of generations. We computed the mean number of secondary cases generated by each case, either based on contact tracing data (using the number of secondary cases observed among traced contacts of each index case), or retrospective data (where cases terminal to the observed chain of transmission were considered to have zero secondary cases). We also fitted a negative binomial distribution to the mean number of secondary cases and estimated the over-dispersion parameter. To deal with secondary cases that had multiple potential infectors, we used a multiple imputation approach: we performed 100 imputations draws, by randomly assigning an infector to these cases, and calculated the pooled mean and 95% confidence interval using Rubin's rules (2). Finally, the serial interval (time interval between symptom onset in infector and infectee) was assessed using the symptom onset dates of (i) pairs identified through contact tracing, and (ii) pairs identified through retrospective investigations. Secondary cases with multiple potential infectors were removed from the serial interval calculations.

**Table S1: Case definition of COVID-19, valid from 17 to 29 January 2020**

| Classification | Definition |
| --- | --- |
| **Possible case** | a) Any patient with clinical signs consistent with severe acute lower respiratory infection requiring admission to hospital with no other etiology that fully explains the clinical presentation AND with a history of travel to or residence in the city of Wuhan, Hubei Province, China, in the 14 days prior to symptom onset.<br>b) Any patient with any acute respiratory illness, whatever the severity, AND with history of at least one of the following exposures in the 14 days prior to illness onset:<br>● close contact with a confirmed case of COVID-2019, while symptomatic;<br>● having shared the same risks of exposure as a confirmed case of COVID-2019 (i.e. same history of travel to or residence in the city of Wuhan, Hubei Province, China);<br>● having worked or attended a health care facility where patients with COVID-2019 have been reported;<br>● having visited or worked in a live animal market in Wuhan, Hubei Province, China.<br>c) Any patient with severe acute respiratory infection for whom an etiology that fully explains the clinical presentation has been initially identified, who develops an unexpected clinical course deterioration AND with a history of travel to or residence in the city of Wuhan, Hubei Province, China, in the 14 days prior to symptom onset. |
| **Confirmed case** | A possible case with a positive SARS-CoV-2 RT-PCR on respiratory samples, performed by an accredited laboratory. |

**Table S2: Case definition of COVID-19, valid from 13 to 30 March 2020**

| Classification | Definition |
|---|---|
| Possible case | a) Any patient with clinical signs consistent with acute respiratory infection with fever or feeling of fever AND with a history of travel to or residence in a risk area, in the 14 days prior to symptom onset.<br>b) Any patient with:<br>● a pneumonia for which another etiology has been excluded based on clinical, radiological and/or virological criteria, and requiring admission to hospital, OR<br>● signs of acute respiratory distress up to ARDS (acute respiratory distress syndrome) in a possibly viral context and without any other obvious etiology. |
| Probable case | Any patient with clinical signs consistent with acute respiratory infection in the 14 days following close contact with a confirmed case. |
| Confirmed case | Any patient, symptomatic or not, with a sample confirming SARS-CoV-2 infection. |

**Table S3: Definition of a contact and follow-up procedure by level of exposure risk, COVID-19, France, January 2020 (adapted from (1)).** Identified contacts were classified into 3 levels of exposure risk: negligible, low, or moderate/high risk, depending on their type of exposure, and different follow-up procedures were subsequently implemented according to the level of risk.

| Level of exposure risk | Contact definition | Follow-up procedure |
|---|---|---|
| Negligible risk | Person who had short (< 15 min) contact with a confirmed case in public settings such as in public transportation, restaurants and shops; healthcare personnel who treated a confirmed case while wearing appropriate PPE* without any breach identified. | Neither identification nor information of contacts. |
| Low risk | Person who had a close (within 1 m) but short (< 15 min) contact with a confirmed case, or a distant (> 1 m) but prolonged contact in public settings, or any contact in private settings that does not match with the moderate/high risk of exposure criteria. | Contacts are asked to measure their body temperature twice a day and check for clinical symptoms. In case of occurrence of symptoms like fever, cough or dyspnoea, contacts are asked to wear a surgical mask, isolate themselves and immediately contact the emergency hotline (SAMU-centre 15) indicating that they are contacts of a confirmed COVID-19 case. |
| Moderate/ high risk | Person who had prolonged (> 15 min) direct face-to-face contact within 1 m with a confirmed case, shared the same hospital room, lived in the same household or shared any leisure or professional activity in close proximity with a confirmed case, or travelled together with a COVID-19 case in any kind of conveyance, without appropriate individual protection equipment. Healthcare personnel who treated a confirmed case without wearing appropriate PPE or with an identified breach. | In addition to the above, contacts are asked to stay at home during a 14-day period after their last contact with the confirmed case while symptomatic and to avoid contacts with the other persons living in the same household (or at least wear a surgical mask). The follow-up consists of an active follow-up through daily calls from the regional follow-up team organized by the Regional Health Agency in collaboration with Santé publique France. |

* PPE: personal protective equipment

**Table S4: Comparison of case characteristics and type of relationships between contact tracing and retrospective investigations.**

| | Contact tracing | | Retrospective investigations | |
|---|---|---|---|---|
| | **Infector** | **Infectee** | **Infector** | **Infectee** |
| **Case characteristics** | | | | |
| Median age, years (IQR) | 51 (37-67) | 48* (30-62) | 47 (34-60) | 54* (46-68) |
| Proportion of children < 15 years old, % (n/N) | 3 (4/127) | 12 (28/236) | 0 (0/24) | 2 (1/57) |
| Proportion of female, % (n/N) | 51 (67/131) | 55 (137/247) | 52 (13/25) | 41 (24/58) |
| **Type of relationships** | | | | |
| Family, % (n/N) | 52 (134/259) | | 42 (29/69) | |
| Coworker/Friend, % (n/N) | 26 (67/259) | | 33 (23/69) | |
| Travel with a case, % (n/N) | 7 (17/259) | | 0 (0/69) | |
| Nosocomial, % (n/N) | 3* (8/259) | | 14* (10/69) | |
| Other/Unknown, % (n/N) | 12 (33/259) | | 10 (7/69) | |

* Differences are statistically significant ($p<0.05$)

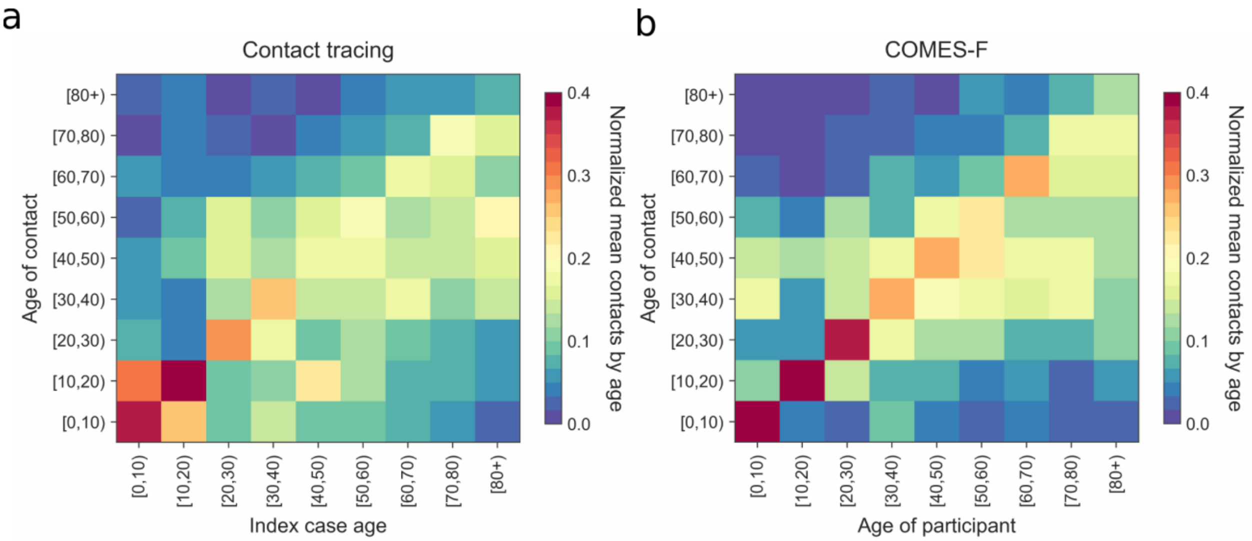


**Figure S1: Contact matrices from contact-tracing data and COMES-F study:** the entries of each contact matrix correspond to the probabilities for each age group to have a contact with another age group. (a) Contact-tracing data. (b) COMES-F study.

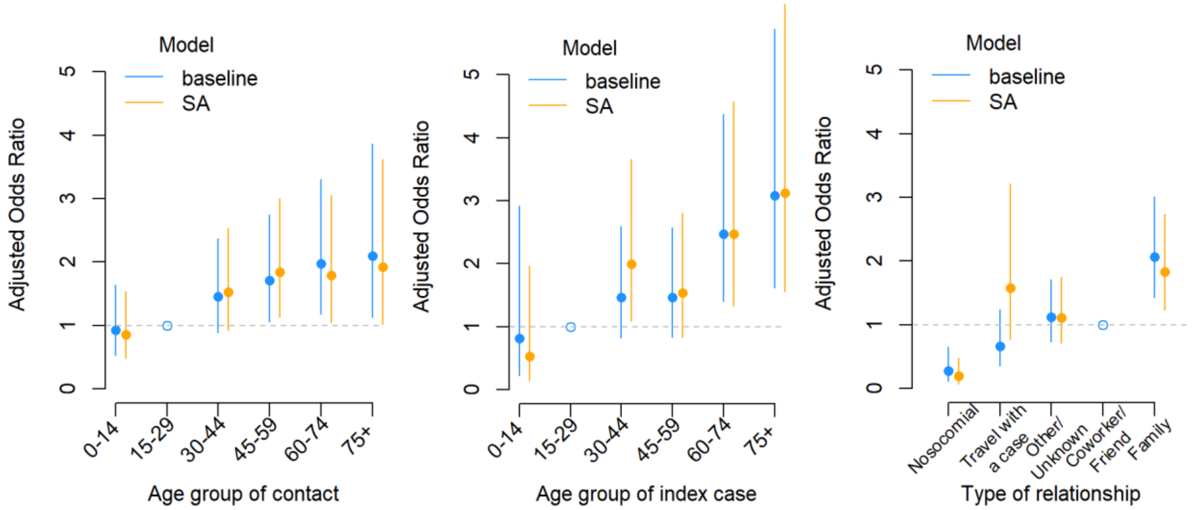


**Figure S2: Results of the sensitivity analysis (SA) including the region of index cases and the time period in the multivariable model.** Adjusted odds ratios of the association between contact becoming a case and contact age (reference: 15-29 years old) (A), index case age (reference: 15-29 years old) (B) and type of relationship (reference coworker/friend) (C). When accounting for regional and temporal differences in data collection, the estimates were not substantially modified compared to the baseline model. The two main differences were an increase in the adjusted odds ratio for contacts whose index cases is 30-44 years old, from 1.5 (95%CI 0.8-2.6) in the baseline analysis to 2.0 (95%CI 1.1-3.7) in the sensitivity analysis, and an increase in the adjusted odds ratio for contacts travelling with a case, from 0.7 (95%CI 0.4-1.2) in the baseline analysis to 1.6 (95%CI 0.8-3.2) in the sensitivity analysis, but confidence intervals were overlapping.

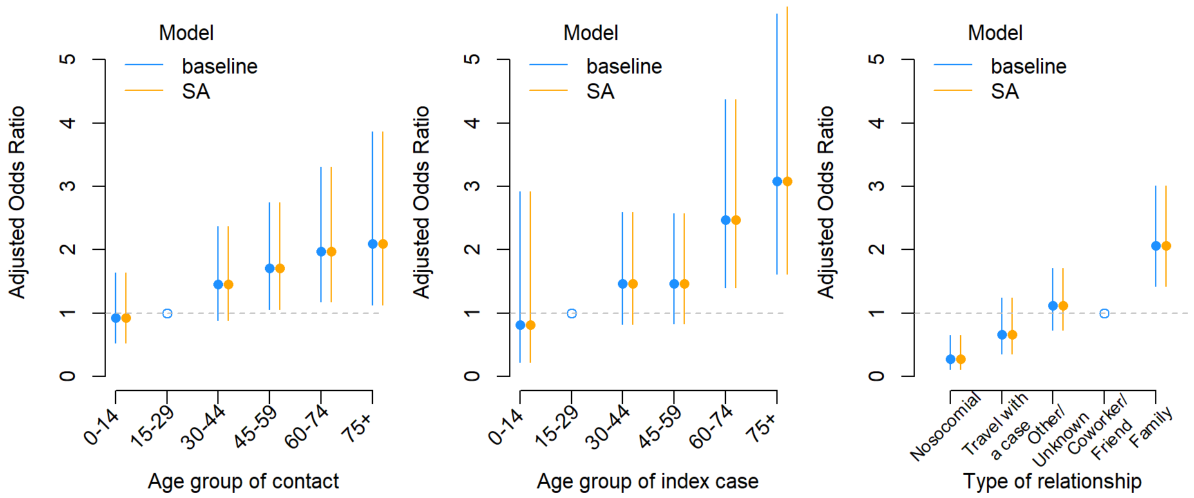


**Figure S3: Results of the sensitivity analysis (SA) restricting the data to moderate/high-risk contacts only.** Adjusted odds ratios of the association between contact becoming a case and contact age (reference: 15-29 years old) (A), index case age (reference: 15-29 years old) (B) and type of relationship (reference coworker/friend) (C). Restricting the analysis to moderate/high-risk contacts did not affect the estimates.

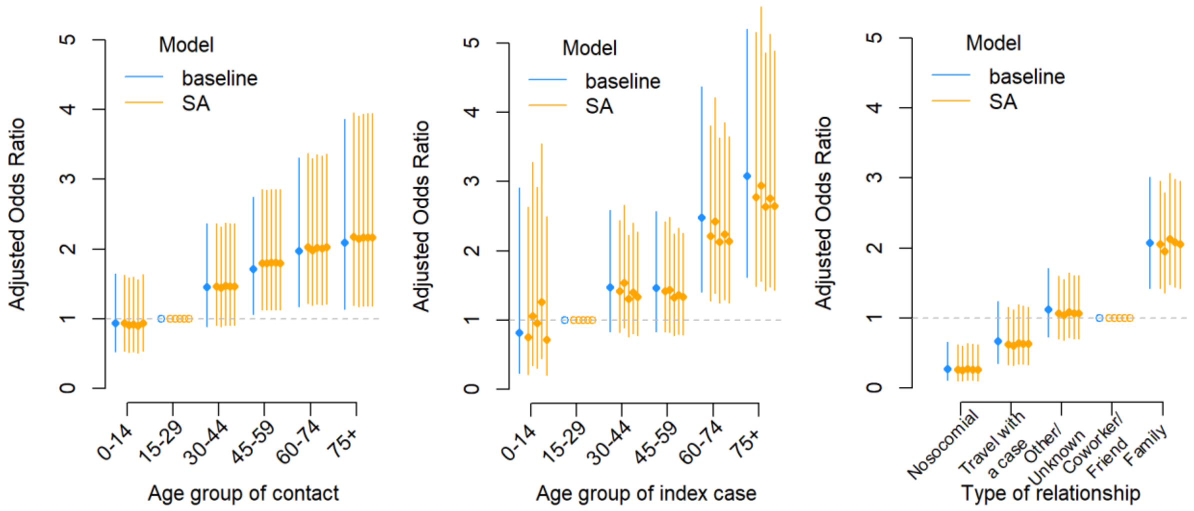


**Figure S4: Results of the sensitivity analysis (SA) on contacts with multiple index cases.** Adjusted odds ratios of the association between contact becoming a case and contact age (reference: 15-29 years old) (A), index case age (reference: 15-29 years old) (B) and type of relationship (reference coworker/friend) (C). Fifty-three contacts had multiple index cases (52 had two index cases and one contact had three index cases). We evaluated the sensitivity of risk factor estimates to inclusion of these contacts by random assignment of a single index case. In the three panels, the five orange points correspond to the estimates obtained on five different datasets, in which one single index case randomly drawn among the multiple index cases of a contact was assigned to this contact. In the five scenarios, the estimates were not substantially modified compared to the baseline model.
